# A NATIONAL SURVEY SHOWS A DILEMMA TOWARDS SBP PROPHYLAXIS AMONG US-BASED HEPATOLOGY PROFESSIONALS

**DOI:** 10.1101/2025.02.24.25322818

**Authors:** Deborah DiazGranados, Jacqueline G. O’Leary, Madhumita Yamuzala, Shari Rogal, Jasmohan S Bajaj

## Abstract

**Background:** Spontaneous bacterial peritonitis prophylaxis (SBPPr)-related practices are evolving, with recent studies showing almost half of potential subjects not being initiated on it.

**Aim:** Determine practice dilemmas regarding SBPPr among US-based hepatology providers.

**Method:** A questionnaire regarding primary and secondary SBPPr using quantitative and qualitative (open-ended) approaches was sent to US-based hepatology providers electronically.

**Results:** 113 clinicians (86% physicians, 73% academic centers) responded. 54% started Primary and 72% secondary SBPPr in 50% of eligible patients. However, the issues related to antimicrobial resistance (AMR) and ineffectiveness lead to SBPPr usage variations and restrictions on a patient-specific basis. Most respondents (>70%) would withdraw/not initiate SBPPr with data regarding ineffectiveness and harms. Open-ended answers showed that most believed newer trials to reduce reliance on weaker older evidence are needed.

**Conclusion:** A survey of US-based hepatologists demonstrates a major dilemma between usual care of SBPPr versus not initiating/withdrawing SBPPr that needs newer randomized trials.

---

Patients with cirrhosis and ascites have a high risk of developing spontaneous bacterial peritonitis (SBP), which is associated with a poor prognosis(1). According to most guideline documents, SBP prevention should be accomplished with long-term antibiotic prophylaxis (SBPPr) to prevent 1st SBP (primary SBPPr) in select patients and recurrent SBP (secondary SBPPr) in all patients(2). However, the data educating these strategies is from trials performed several decades ago(2-4). Recently, due to increasing antimicrobial resistance (AMR), reports of both primary and secondary SBPPr ineffectiveness and potential harm have emerged leading to clinical care dilemmas (5-8). Moreover, recent reports show that a sizable proportion of patients do not receive this as usual care(9). However, the attitudes of US-based clinicians and practice differences regarding SBPPr need further elucidation. We aimed to examine clinician attitudes across the US regarding SBPPr using a standardized survey.

## Method

With input from social science colleagues (DDG and MY), we designed an online survey that inquired about demographics, practice details, awareness of AMR, and current practice regarding both primary and secondary SBPPr. Additionally, questions pertaining to dilemmas in current management of SBPPr were posed. We also allowed open-ended comments regarding current SBPPr practices. The questionnaire was sent using REDCAP after approval by Richmond VA Research and Development Committee and VA Employee Union to hepatology providers (both physician and midlevel) in VA Central leadership, their university affiliates, and leading US-based hepatologists. The questionnaire was sent between November 2024 and January 2025 with two reminders, and results were collected in a de-identified manner. Responses were collated and compared for primary and secondary SBPPr and summarized.

## Results

A total of 113 participants responded to our survey, results report participants who provided responses for each question; not all 113 participants responded to every question. Demographics: 86% were physicians and the remaining were nurse practitioners. Most were from academic centers (73%), 21% VA, 3% both and remaining 2% from private centers. Most respondents (58%) were in practice for >11 years, reported they saw >25 (63%) patients a month with decompensated cirrhosis and 54% of respondents started Primary SBPPr and 72% of respondents reported starting secondary SBPPr in >50% of their eligible patients.

### AMR

As shown in table 1, most respondents believed that AMR was problematic and compounded by antibiotic prescribing practices in primary care, inpatient and hepatology practitioners. Respondents thought that patients with cirrhosis did not expect nor would demand SBPPr since this is mostly a clinician decision.

**Table 1:**
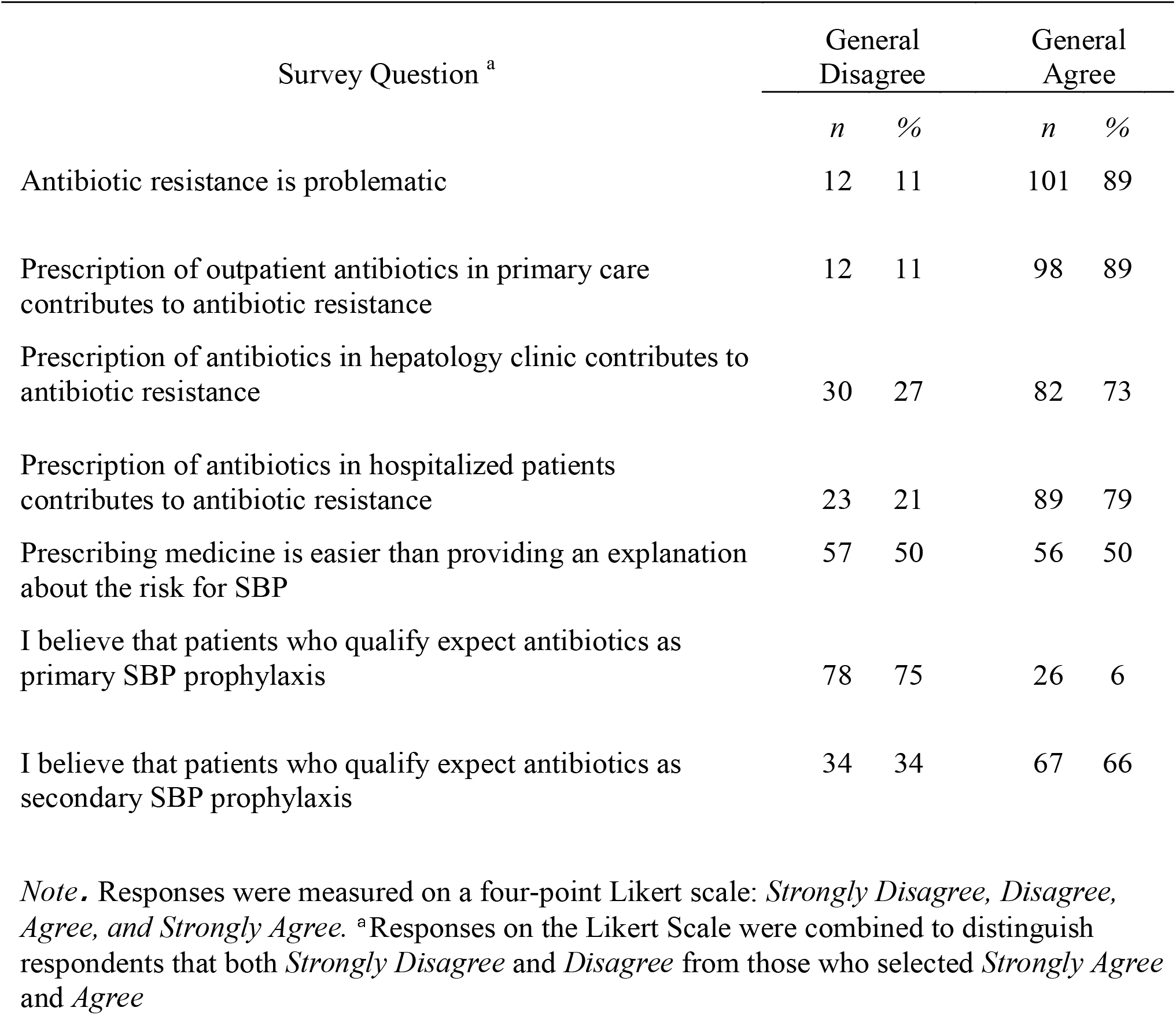
Responses to questions regarding status of SBP prophylaxis.

### Dilemma regarding current state of SBPPr: Primary SBPPr

66% felt there was adequate data regarding the ineffectiveness of primary SBP to prevent the first episode of 1^st^ SBP. 76% acknowledged a sufficient lack of mortality benefit and 66% all-cause hospitalizations to stop SBPPr. Importantly, 80% agreed there was potential for harm, 64% including adverse events, contributing to the dilemma of primary SBPPr initiation. If further evidence was presented up to 70% would not initiate or would withdraw Primary SBPPr. Secondary SBPPr: 73% felt there was data regarding ineffectiveness in preventing SBP, and 79% a lack of mortality benefit, including 72% all-cause hospitalizations. Importantly, 81% felt there was potential for harm (similar to primary SBPPr), including 64% felt adverse events contributed to this dilemma. If further evidence was presented up to 72% would not initiate or would withdraw secondary SBPPr.

### Rationale and dilemma resolution using open-ended questions

Most respondents said they still initiated secondary SBPPr (65%), but fewer were initiating primary SBPPr (45%) because of guidelines, but they acknowledged these guidelines were potentially outdated. Some respondents preferentially used SBPPr in transplant candidates, those with high MELD scores, and those with multiple admissions. However, they acknowledged that the practice of picking and choosing whom to start on SBPPr was not guideline driven and needed resolution. Most respondents called for new prospective randomized clinical trials, updated evidence from systematic reviews, meta-analyses, and newer guidelines with educational material (Table 2).

**Table 2:**
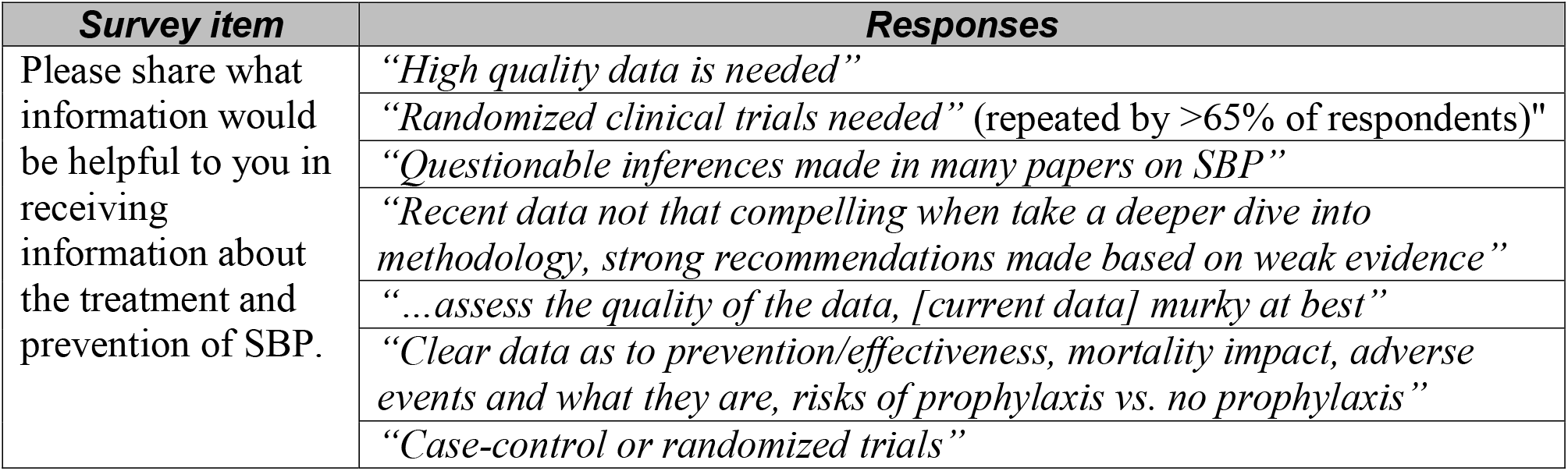
Themes of responses to open-ended question regarding status of SBP prophylaxis.

| **Survey item** | **Responses** |
| --- | --- |
| Please share what information would be helpful to you in receiving information about the treatment and prevention of SBP. | *“High quality data is needed”* |
|  | *“Randomized clinical trials needed”* (repeated by >65% of respondents)" |
|  | *“Questionable inferences made in many papers on SBP”* |
|  | *“Recent data not that compelling when take a deeper dive into methodology, strong recommendations made based on weak evidence”* |
|  | *“...assess the quality of the data, [current data] murky at best”* |
|  | *“Clear data as to prevention/effectiveness, mortality impact, adverse events and what they are, risks of prophylaxis vs. no prophylaxis”* |
|  | *“Case-control or randomized trials”* |

## Discussion

In this national survey of US-based hepatology providers, there was marked skepticism regarding the utility of both primary and secondary SBPPr, which encapsulates the dilemma for clinicians.

The survey touched on salient points regarding primary and secondary SBPPr, which have differing risk profiles and patient burdens(10, 11). Primary SBPPr has less robust evidence and, of late, is associated with higher AMR among those who later developed SBP(10, 12). With increasing AMR and individual practice variations, there were even major differences in secondary SBPPr in recent national studies. Almost half of patients eligible for secondary SBPPr were not initiated on it in both Veteran and non-Veteran cohorts (9). This dilemma was further confirmed by our survey where substantial variations among practitioners was evident with some reserving it for more advanced patients or transplant listed patients and other individualized approaches. Furthermore, respondents were willing to not initiate and even withdraw SBPPr in patients with ascites if newer data showed ineffectiveness or harm and called for prospective trials to resolve the dilemma. The equipoise among practitioners towards use of SBPPr was also found in a recent Cochrane systematic review(5). Issues related to unnecessary antibiotics include AMR, adverse effects of the antibiotics, and lack of efficacy while the opposing side of the decision-making process includes preventing SBP, which remains a serious complication(13).

The data could be biased since surveys were sent to national VA affiliated individuals and their university colleagues. However, most respondents are affiliated with universities, increasing the generalizability.

We conclude from our survey of hepatology providers that there is a dilemma regarding long-term antibiotic prophylaxis to prevent SBP occurrence and recurrence. Those who continued to initiate SBPPr cited guidelines, although the most recent guidance document did not recommend primary SBPPr(14). While others either do not use it due to AMR or restrict it to specific groups of patients, adding to the dilemma in clinical practice. There is a need for future trials to determine which practice pattern of SBPPr; initiation and/or continuation versus no initiation and/or discontinuation, is more effective in reducing SBP occurrence and recurrence. The survey respondents indicated willingness to change practice including not initiating or withdrawing SBPPr if newer evidence shows the practice is ineffective or harmful.

## Data Availability

Individual survey data are not available

## Acknowledgements

We acknowledge Ms Sandra Gibson for assistance with obtaining VA Labor Union approval for distributing the questionnaire.

